# Fractional-Order SEIHR(D) Model for Nipah Virus with Spillover: Well-Posedness, Ulam–Hyers Stability, and Global Sensitivity

**DOI:** 10.64898/2026.02.02.26345408

**Authors:** Taylan Demir, Hasan Hüseyin Tosunoğlu

## Abstract

In this research, we create a new fractional-order SEIHRD framework to examine how the Nipah virus moves from one species to another (zoonotic spillover) and how it later spreads throughout a community (via contact with one another) or in a hospital or isolation situation (via entering into a hospital or being placed under quarantine). We used the fractional-derivative formulation of the SEIHRD model to demonstrate memory-based effects related to the progression of an infection and also reflect time-distributed effects associated with surveillance and control measures placed on an infected patient. We first demonstrated that the basic epidemiologic properties of the model were consistent by showing that the solutions of the SEIHRD differential equations will always yield positive and bounded solutions within biologically relevant parameter ranges. We then established the well-posedness of this model by transforming the SEIHRD differential equations into an equivalent integral operator and applying various fixed-point arguments to demonstrate that there will always be unique solution(s) to the SEIHRD differential equations. To evaluate the threshold parameter for the transmission of Nipah virus within a given population we calculated the threshold level through the next generation method to determine the expected number of secondary infections from a new or chronically infected host. One of the main contributions of this work is to include an analysis of the robustness of a given solution to all potential perturbations (i.e., Ulam-Hyers and generalized Ulam-Hyers stability). In addition, we provide analytic results guaranteeing that small perturbations due to approximate modeling, numerical approximation (discretization), or the lack of data fidelity will produce controlled deviations in the solutions. To finish this project, we perform a global sensitivity analysis on uncertain coefficients to evaluate their contribution to the uncertainty of each coefficient and to find out the coefficients that most strongly influence major outcome metrics. This will allow us to develop a priority order for prioritizing spillover control (reduction of human contact and/or isolation), contact reduction, and expenditure of resources towards isolation-related interventions. The resulting framework converts fractional epidemic modeling from a descriptive simulation to a replicable method with robustly defined behavior and equal response prediction.

## 1 Introduction

The Nipah virus is one of several zoonotic (animal to human) diseases threatening our global health, as these pathogens do not discriminate when evolving in wildlife and humans. Fruit bats belonging to Pteropus are a natural reservoir for the Nipah virus. Most human cases occur due to spillover from wildlife via movement patterns and the way food is handled in restaurants. For example, there have been numerous contamination events tied to bat excrement (urine, feces) prior to or during close contact with the infected person through various means of providing care. Additionally, there are hospitals that have reported contaminated surfaces due to bat urine or feces that have led to transmission of the virus to patients in these health care facilities [2]. Public health is not only about documenting the transmission of an outbreak once it has already started, but also about learning how spillover from wildlife creates new cases of disease in humans, and how these cases may subsequently be transmitted between humans. While there are many challenges to accomplish this with incomplete information, delayed identification and isolation of individuals with the infection, and variability in patterns of contact between people, understanding the relationship between these factors is crucial in addressing the public health challenges of Nipah virus and other related henipaviruses. This, along with other public health needs, led to Nipah and other related henipaviruses being established as a priority for urgent research and development initiatives as part of global preparedness planning. In many areas, prevention and supportive care are currently the primary methods of controlling outbreaks [3,12,13]. Mathematical models are vital for capturing the complexity of these types of interactions; however, Nipah uniquely poses a modeling conflict on two levels: as an external force (as spillover) that enters into an infection rate in humans, while also being commonly defined by limited/unstable human-to-human transmission rates reliant on the particular context in which transmission occurs (e.g., caregiving in the household, close contact with infected individuals). The data-driven investigations of these issues carried out in Bangladesh have demonstrated the involvement of both contaminated date palm sap pathways (as a route for viral transmission) and the role that close proximity of infected individuals to uninfected individuals plays in transmitting virus into the general population; therefore, prevention should consist of both reducing exposure to infected individuals and preventing contact-mediated viral spread [1,6]. Thus, in order for a model to be operationally relevant, it should (i) account for the spillover effect in a manner that is explicit rather than implicit within a single effective rate of transmission, and (ii) retain interpretability of key results such as threshold properties associated with secondary transmissions, outbreak size, time of peak, and cumulative severe cases in a manner that is transparent to both mathematical and public health audiences [9]. An additional complication associated with epidemic trajectories is that they exhibit “memory” in multiple nontrivial ways: the progression of infection over time, delays in reporting data, behavioural adaptation, and control measures will all create temporal effects that will not necessarily be well captured by local-in-time dynamics alone. By replacing the traditional derivative with a fractional operator, fractional-order formulations provide a systematic method for encoding non-local dependence in the dynamics of infectious disease (epidemic) processes so that the behaviour of the present state can be influenced by a historically weighty state. Recent research on fractional epidemic models has suggested that this framework provides greater descriptive flexibility than currently available empirical evidence and may aid in the interpretation of some forms of memory-like effects if appropriately justified and validated through independent experimentation [9]. A fractional compartmental model offers an effective way of mathematically joining the historical exposure to Nipah with the current risk of transmission by providing an agreed upon framework through which to understand how both early and later surveillance and isolation efforts may have influenced the course of activity in the Nipah environment. We present a fractional-order SEIHRD-type model to investigate spillover as an explicit seeding method into the exposed class, community transmission as well as healthcare-associated dynamics represented through immediately interpretable interaction terms and by including a specific compartment to capture hospitalization or isolation. The analysis meets the highest standards of mathematical and epidemiological reliability through the establishment of well-posedness in a biologically meaningful domain, via proving positivity and boundedness as well as finding existence and uniqueness through fixed-point methods applied to an equivalent integral equation. We derive a reproduction metric by using the next-generation method to quantify threshold behavior so that we can provide an accurate description of the secondary transmission potential from an importation-based action due to spillover. Thirdly, and most important for the robustness of the proposed framework, we prove Ulam-Hyers and generalized Ulam-Hyers stability for fractional systems, thereby proving analytically that approximate solutions obtained through perturbations arising from modeling errors, discretization errors, or errors arising from data, etc., are always uniformly close to the exact solution, provided certain explicit conditions are met. The initial concept of stability was introduced by Hyers, and it has been extended using a variety of approaches, including those used for fractional differential systems [4,5,8,10]. To be able to use mathematical models to gain useful insights, we conduct multiple global uncertainty and sensitivity analyses, using standard sampling-based and variance-based approaches (e.g. PRCC and Latin-hypercube sampling) to determine the influence of all parameters on each of the critical system outputs in the presence of many uncertainties [7,11]. The rest of this document provides an explicit framework for understanding the theory/computation/interpretation pipeline through four stages. In the first stage, we create a fractional modeling framework and provide the definitions necessary to explain it. The second stage, model foundation and threshold characterization, will provide evidence of the validity of the model methodology. The third stage, the development of Ulam-Hyers stability, will document development of Ulam-Hyers stability for the proposed Nipah dynamics. The fourth stage, the application of uncertainty and sensitivity techniques, will provide numerical experiments aimed at illustrating trajectories, validating theoretical bounds, ranking the relative importance of different intervention parameters under conditions of uncertainty. We will end by discussing the implications of these results regarding reducing transmission via spillover, reducing contact with infected animals or humans, and how effective isolation can reduce transmission from previous contacts. Finally, we will briefly discuss extensions necessary to account for substantially greater degrees of heterogeneity and ways to conduct datadriven calibration.

## 2 Preliminaries and Notation

Throughout the paper, let *T* > 0 be fixed and consider all state variables on the time interval [0, *T*]. For a given integer *m ≥* 1, we work in the Banach space *C*([0, *T*]; **R**^*m*^) equipped with the uniform norm ||*x*||_∞_ = *sup*_*t*∈[0,*T*]_ ||*x*(*t*)||, where ||.|| denotes the Euclidean norm in **R**^*m*^. When a positively invariant set Ω ⊂ **R**^*m*^ is specified, we interpret the nonlinear vector field *F* : [0, *T*] × Ω → **R**^*m*^ as acting on trajectories *x*(.) ∈ *C*([0, *T*]; Ω). A standing assumption used in the well-posedness and stability arguments is a Lipschitz condition in the state variable: there exists *L* > 0 such that ||*F* (*t, x*) − *F* (*t, y*)|| ≤ *L* ||*x* − *y*|| for all *t* ∈ [0, *T*] and all *x, y* ∈ Ω. This setting is standard for fractional initial-value problems and is particularly convenient when one aims to obtain explicit robustness bounds.

### 2.1 Caputo fractional derivative and integral equivalence

We recall the Caputo fractional derivative of order *α* ∈ (0, 1), which is the natural choice when the initial conditions are prescribed in the classical (integer-order) sense. For a function *x* that is absolutely continuous on [0, *T*], its Caputo derivative is defined by

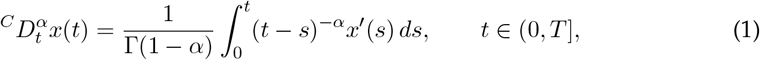

where Γ(·) denotes the Gamma function [14-16]. The associated Riemann–Liouville fractional integral of order *α* > 0 is given by

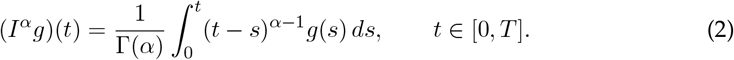

One significant technical point applicable to future work is that a fractional differential equation of the Caputo type can be transformed into an integral equation with Volterra-type kernel. We will begin by defining the fractional initial value problem (IVP) defined next

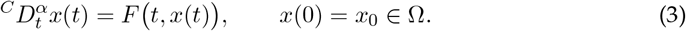

With some mild assumptions on regularity conditions on F, (3) is equivalent to using

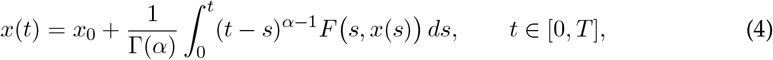

as the starting point of the fixed-point approach presented in next section and of the quantitative estimates established in reference [14-16] for Ulam-Hyers stability. In particular, once the dynamics are expressed in integral form, we can make use of Lipschitz continuity and appropriate fractional integral inequalities to provide a means for controlling the differences in solutions.

### 2.2 Auxiliary results and the fixed-point framework

The integral formulation suggests a natural operator *T* acting on *C*([0, *T*]; ℝ^*m*^) via

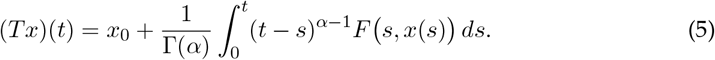

When *F* is Lipschitz on Ω with constant *L*, one obtains the estimate

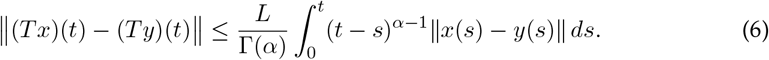

Taking the supremum over *t* ∈ [0, *T*] yields a convenient sufficient condition ensuring that *T* is a contraction on a suitable closed subset of *C*([0, *T*]; ℝ^*m*^), which leads to existence and uniqueness by a standard fixed-point argument [16]. If contraction is not ensured on all intervals (or for the whole parameter range), we can utilize compactness-based alternatives to establish a priori bounds. This usually means that we can show that *T* is continuous and takes a bounded closed convex set to a relatively compact set. In this work, we focus on assumptions that are verifiable for the epidemiological vector field and where the constants involved can be kept as explicit in the robustness bounds of 2.3 since these constants reappear as we stated above. A second ingredient repeatedly used in stability estimates is a fractional Grö nwall-type inequality. Let *u* ∈ *C*([0, *T*]) be nonnegative and suppose that, for some constants *a, b ≥* 0, it satisfies

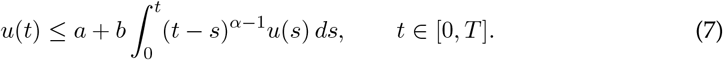

Then *u* admits an explicit bound in terms of the Mittag–Leffler function *E*_*α*_, namely

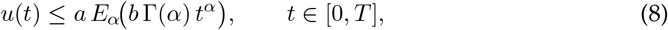

which is well adapted to Caputo-driven dynamics and provides a sharp tool for turning integral inequalities into uniform-in-time estimates [17]. The significance of estimating robustness bounds stems from the fact that we can quantify the spread of perturbations through trajectories and represent the final robustness constants in relation to *α, T*, and the Lipschitz constant, *L*.

### 2.3 Ulam-Hyers stability concept for fractional systems

The Ulam-Hyers stability perspective gives formalism to the idea of a robustness principle, i.e., that if the trajectory of a system is behaved according to the governing dynamics, but is only approximately so due to perturbations in models (e.g., due to numerical discretization), inaccuracies in the data, etc. then the trajectory must remain close (uniformly) to some exact solution of the system (in the sense of trajectory). The ideas that underlie this perspective originated in Ulam’s original question about stability, which was answered by Hyers [4] in an early work in the classical case, and these same ideas have been generalized greatly in the Ulam-Hyers-Rassias direction [10]. The Ulam-Hyers-Rassias ideas have been adapted for use in the case of fractional differential equations within the last several years, where non-locality makes it particularly valuable to obtain explicit error bounds [8].

In our Caputo setting, let *y* ∈ *C*([0, *T*]; Ω) be an approximate solution of

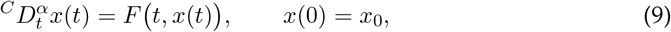

in the sense that there exists *ε* > 0 such that

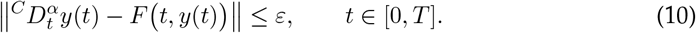

On the interval [0, *T*], it is called Ulam-Hyers stable if there exists a constant *C* > 0, independent of *ε* and of the approximate solution *y* in question, such that there are exact solutions *x* to the model satisfying

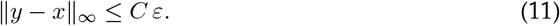

A frequently used extension is *generalized Ulam–Hyers stability*, where the linear control *Cε* is replaced by a nondecreasing function *ϕ*(*ε*) with *ϕ*(0) = 0, yielding

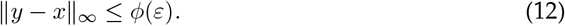

These definitions are a way of describing the fact that if there is a bounded defect in the differential law, there will be an associated bounded deviation in the explicitly defined trajectory of the solution in the uniform norm. The estimates in the following sections are obtained from using an integral form (because this uses all available information), applying the Lipschitz type property of *F*, followed by closing the resulting inequality via the fractional Grö nwall case. The result of all this leads to constants which depend on *α, T*, and *L* [17]. The close connection between the approximate dynamics of the epidemic systems and uniform closeness is what renders Ulam–Hyers stability a valid analytical certificate of the robustness of the simulated and data-driven epidemic models [4,8,10].

## 3 Model Formulation

### 3.1 Compartments and biological assumptions

We examine a population of humans living with Nipah virus transmission through animal hosts. The virus can be transmitted from humans when there are unintentional interactions between species. Based on current epidemiology data, spillovers will happen when humans come into contact with a natural reservoir for Nipah, namely the Pteropus bat. Spillovers will also happen when humans consume food contaminated with Nipah or their symptoms by administering date palm sap, which is also linked to transmission from animals to people. In our model, we have represented spillover as separate from the total contacts that may transmit, therefore using two separate means of estimating the spillover to people [1,6,12]. The population will be divided into five areas that capture the entire population for use in epidemiological modelling: Susceptible (*S*(*t*)); Exposed (*E*(*t*), i.e. infected but not yet infectious); Infectious (*I*(*t*), i.e. infectious to the local population); Hospitalized/Isolated (*H*(*t*)); and Recovered (*R*(*t*)). The cumulative number of deaths as a result of illness is tracked by *D*(*t*). Therefore, *N*(*t*), or the living population size, can be described as the sum of the other five compartments as follows: *N*(*t*) = *S*(*t*) + *E*(*t*) + *I*(*t*) + *H*(*t*) + *R*(*t*) and does not include *D*(*t*) because *D*(*t*) records only mortality due to illness. Separating mortality due to the disease from the total population is beneficial because it allows *D*(*t*) to be considered a public health outcome, free of any other considerations related to the balance of the living population. The force of infection (the frequency dependence) for the community is based on the effective contact rate between different population classes. This means that when an outbreak occurs, the scale of infection depends on how well populations mix as opposed to their actual size (density). We define

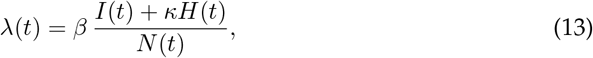

where *β* > 0 is the effective transmission coefficient and *κ* ∈ (0, 1) encodes reduced infectiousness of hospitalized/isolated individuals due to infection-control measures (e.g. barrier nursing, restricted contact, or dedicated wards). As *H*(*t*) appears in (13) it indicates that healthcare settings may contribute to disease transmission when control measures are not fully effective. Documentation exists of hospital surfaces with Nipah virus contamination during Nipah epidemic investigations in 2002-2003 [2]. The requirement that *κ* < 1 supports biological evidence that isolation decreases the rate of ongoing disease transmission relative to the amount of ongoing infectiousness in a community that is not isolation. The introduction of spillover through a nonnegative fuction, *η*(*t*), is intended to model the way in which susceptibility can serve as a seeding mechanism (origin/semi-origin) for the entire exposed population. Multiple possible hypotheses can be modeled in this formulation.(i) a constant baseline spillover *η*(*t*) = *η*_0_; (ii) seasonal modulation consistent with ecological variation, for instance 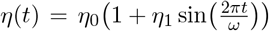 with |*η*|_1_ < 1; or (iii) short episodic bursts representing discrete exposure events (approximated by narrow pulses rather than idealized distributions). These options enable the model to identify distinctions between “importation pressure” and the internal transmissibility of disease. This distinction is vital to interpreting thresholds and points at which to intervene [12,13].

### 3.2 Full fractional-order system (Caputo *α*)

The time delay that can be caused by non-instantaneous infection progression, reporting delays and behavioral adaptation, lagged effects of interventions, will be captured in the model by the use of Caputo fractional derivatives of order *α* ∈ (0, 1]. The case of *α* = 1 will return standard (or classical) integers, whereas fractional-order formulations are becoming a standard companion for model development of epidemics when the goal of the analysis is to provide parsimony and to account for the long-term dependency of the history of the epidemic and its effects [9]. Therefore, the parameters in the model will have their values interpreted in terms of the time-scale being used and their units in relationship to that time-scale as well as in reference to the Caputo operator in a fractional manner.

Let *S*(*t*), *E*(*t*), *I*(*t*), *H*(*t*), *R*(*t*), and *D*(*t*) denote the susceptible, exposed, infectious, hospitalized/isolated, recovered, and cumulative death classes, respectively, and let

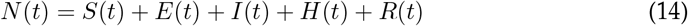

be the living population size. The force of infection is given by

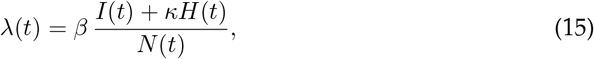

where *β* > 0 is the effective transmission coefficient and *κ* ∈ (0, 1) represents reduced infectiousness under hospitalization/isolation (infection control). The non-negative function *η*(*t*) for measuring the external effect of coalescence on a population in *E*(*t*) can be defined as an outside generation intensity that produces transfers of individuals from *S*(*t*) to *E*(*t*). The proposed Caputo fractional-order SEIHRD model is then defined by;

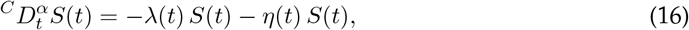

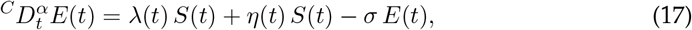

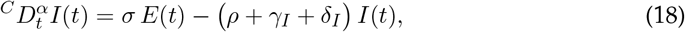

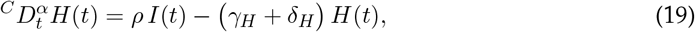

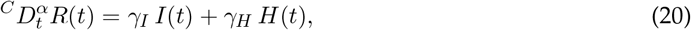

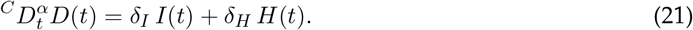

Here *σ* > 0 is the progression rate from exposed to infectious, *ρ* > 0 is the hospitalization/isolation rate, *γ*_*I*_, *γ*_*H*_ ≥ 0 are recovery rates, and *δ*_*I*_, *δ*_*H*_ ≥ 0 are disease-induced mortality rates in the infectious and hospitalized classes, respectively.

The initial conditions are prescribed in the classical sense:

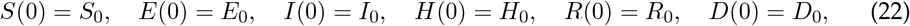

with *S*_0_ + *E*_0_ + *I*_0_ + *H*_0_ + *R*_0_ = *N*_0_ and typically *D*_0_ = 0. To ensure that equation (22) retains its traditional epidemiological meaning as stated in [16], it is necessary to employ the Caputo derivative for the solution. The modeling case is addressed with two comments. The spillover component *η*(*t*)*S*(*t*) quantifies the introduction of expanded exogenous zoonosis and does not replace the transmission of the Nipah virus from human-to-human, instead it differentiates between importation forces and internal transmissibility consistent with the zoonotic nature of the Nipah virus [1,6,12]. In addition, including the term, *H*(*t*) in equation (15) indicates that healthcare-associated transmission can occur when infection control is not successful, while *κ* < 1 indicates a decrease in transmission while in isolation [2].

### 3.3 Parameter definitions and principled selection of ranges

When dealing with datasets about Nipah viruses, it is essential to account for uncertainty in the parameters used to model outbreaks due to the localized nature of outbreaks and sporadic data availability. To deal explicitly and conservatively with this uncertainty, each parameter has been chosen according to the following criteria: (a) the choice of each parameter is made based on a clear understanding of the biological interpretation associated with that parameter (e.g., incubation/latency periods, outcome times); (b) the ability to associate each parameter to a time scale provides a fractional time-scale interpretation consistent with the fractional mathematical dynamics framework; and (c) the selection of the range of each parameter is based on clear, principled criteria for establishing range limits. We recommend selecting ranges based on three criteria: (1) the plausibility window of clinical and epidemiologic evidence reported from multiple sources (i.e., public health [3,12]); (2) evidence of outbreak-related transmission dynamics and routes of contact-mediated spread (including spillover and human-to-human); and (3) a conservative boundary that covers plausible variation (where published data indicate that variation exists among studies) in a variety of epidemiological settings. In performing sensitivity/uncertainty analyses, the wider the defensible range used in the analysis, the more reasonable the “nominal” parameter choices; this reduces the likelihood of an over-narrowing “estimate” of a parameter providing spurious indications of identifiability. Best practices require that justification of each parameter range uses a compromise between available empirical evidence and the use of precautionary principles of uncertainty, especially in instances where estimating parameter details may have major implications on the subsequent ranking or estimate of those parameters [7,11]. In terms of spillover, *η*(*t*) is not treated as just one constant value, instead it functions as an input which allows for scenario analyses to run explicitly: low persistent spillover or a burst type of spillover which would correspond to short period of time clusters. This makes sense because spillover is based on both ecological factors and human behaviours which can sometimes have a longstanding effect. These factors can also change dramatically over time within a given location. [12,13] The hospital-related reduction factor; *κ* will be assumed to have a value from 0-1 and be interpreted based on the effectiveness of controlling infections. Therefore, *κ* would have a closer total outside of the isolation of an infected person than if it was closer to 0 which would mean almost complete isolation, whereas *κ* closer to 1 indicates substantial residua transmission within care settings and *κ* ∈ (0, 1), [2].

### 3.4 Outputs and performance metrics

In order to make the model relevant to identifiable endpoints of outbreaks, we highlight several model outputs that are of direct relevance to public health decision-making. First, the internal transmission threshold is given by the basic reproduction metric *R*_0_, which is calculated using the next generation matrix approach on the infected subsystem, centered at the disease-free state, with the external spillover set to zero, i.e., *η*(*t*) ≡ 0. This is standard when the model under consideration has external transmissions [18,19]. Second, we track peak intensity and timing metrics such as

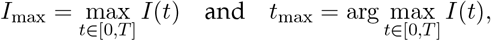

which reflect acute burden and resource planning needs. Third, cumulative mortality *D*(*t*) is employed as a measure for severe outcomes, in line with the severity of outcomes emphasized by public health agencies [12,13]. Finally, we report attack-rate–type summaries such as

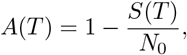

and cumulative-incidence proxies derived from integrated inflows into *E*(*t*). These metrics offer a compact performance layer that can be used to compare spillover scenarios, hypotheses regarding interventions through *κ* and *ρ*, as well as memory effects through *α*.

## 4 Basic Properties: Positivity, Boundedness, Invariant Region

### 4.1 Positivity

The premise of a viable condition of models for compartmental, epidemic will provide a minimum reliability standard— the results must be nonnegative and biologically valued. Given that the model starts with some form of nonnegative data (*S*_0_, *E*_0_, *I*_0_, *H*_0_, *R*_0_, *D*_0_ *≥* 0) it must also remain in a nonnegative state at all times *t* ∈ [0, *T*]. The Volterra-like integral representation related to (*t* − *s*)^*α*−1^ ensures the model’s positivity. *α* ∈ (0, 1) guarantees that the kernel in the integral representation will be positive over [0, *T*]. This property propagates through the model when the right hand side is quasi-positive in the conventional compartmental sense [14,15,16].

To make the argument concrete, note that the susceptible component satisfies an integral relation of the form

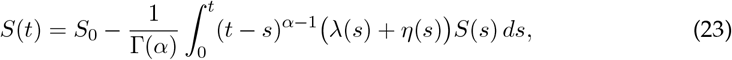

where *λ*(*t*) *≥* 0 is the force of infection and *η*(*t*) *≥* 0 is the spillover input. If the value of *S*(*s*) *≥* 0, then it can be assumed that the value of the integrand will also be nonnegative and as such cannot cause *S*(*t*) to reach a negative value after finite time. Instead, it will contribute a nonnegative depletion to the value of *S*. Similar arguments regarding boundaries apply to the other compartments: every time *E*(*t*) = 0, both (i) the outflow − *σE*(*t*); and (ii) the inflows *λ*(*t*)*S*(*t*) and *η*(*t*)*S*(*t*) will be non-zero; every time *I*(*t*) = 0, (i) the inflow *σE*(*t*) will be non-zero but both (ii) the linear outflows will be zero; and the same will hold true for *H*(*t*). The recovery and cumulative life-death variable will only allow positive inflows *γ*_*I*_*I* + *γ*_*H*_*H* and *δ*_*I*_*I* + *δ*_*H*_*H* therefore no negative outflow from these variables. Since *F* is assumed to have continuous properties and the parameters are assumed to be nonnegative, it is guaranteed that the solution path stays in 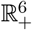 over the time interval from [0, *T*] [14,15,16].

### 4.2 Boundedness and an invariant region

Beyond positivity, the model must remain bounded in a biologically interpretable region. Define the living population

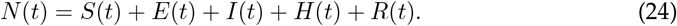

Using the properties of the Caputo operator and summing up the first five equations of the model together results in all of the internal transfer terms cancelling out; thus, the only net loss from the living population results from disease-induced deaths, which are accounted for in *D*(*t*). As a result, the combined quantity *N*(*t*) + *D*(*t*) satisfies

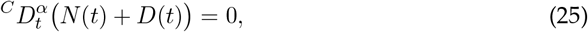

and hence

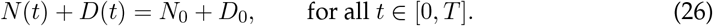

Specifically when *D*_0_ = 0 and *N*(*t*) + *D*(*t*) = *N*_0_ demonstrating the individuals in the model have closed accounting records. As such, this identity has two immediate implications: It must be true that *D*(*t*) is non-decreasing; its integral representation is:

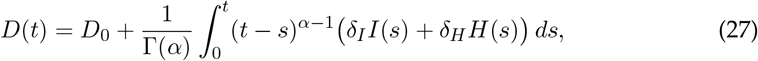

has a nonnegative integrand, and the kernel is nonnegative, implying that *D*(*t*) cannot decrease with time. Moreover, it follows from the preservation of the current level of biodiversity that 0 ≤ *D*(*t*) ≤ *N*_0_ + *D*_0_ and 0 ≤ *N*(*t*) ≤ *N*_0_ + *D*_0_ for all *t* in the interval [0, *T*]. Therefore, we can assume that the natural set of all biologically meaningful measurements is;

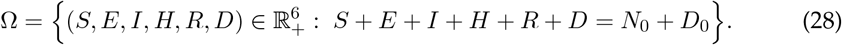

Together, positivity and the conservation law imply that all trajectories beginning in Ω will remain in Ω for all *t* ∈ [0, *T*]. Furthermore, since Ω is compact (for well-defined values of *N*_0_ + *D*_0_), it gives a natural domain in which to define global Lipschitz or one-sided growth conditions on which we can base our subsequent arguments of well-posedness and robustness [14,15,16].

### 4.3 Continuous dependence on initial data and parameters

Lastly, one final, but very important, property that connects the analysis of the result to numerical simulations, as well as to Ulam-Hyers stability, is the dependence of the solution on the choice of initial data and parameters being continuous. Writing the system in integral form,

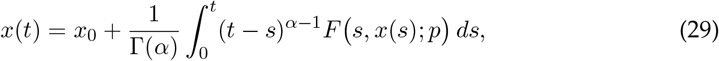

where *p* denotes the parameter vector, one may compare two solutions *x*(·) and 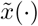 associated with (*x*_0_, *p*) and 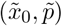. Under Lipschitz continuity of *F* in the state variable on Ω, and mild regularity with respect to parameters, one obtains an inequality of the form

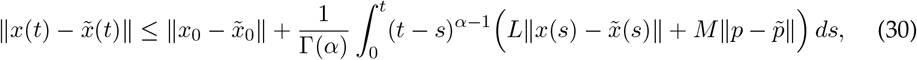

for suitable constants *L, M ≥* 0. Closing this inequality with a fractional Grö nwall estimate yields a uniform bound in terms of the Mittag–Leffler function *E*_*α*_, typically of the form

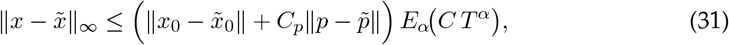

with constants depending only on *α, T*, and the Lipschitz-type bounds on *F* [17]. This result fulfills two types: providing stability of the solution operator, thus completing the well-posedness picture, and providing an exact mechanism through which small perturbations (due to data uncertainty, discretization or modeling error) will cause bounded deviations of the trajectories - this is the exact conceptual underpinning of the Ulam-Hyers framework developed later on. [16,17].

## 5 Well-Posedness: Existence and Uniqueness

### 5.1 Construction of the integral operator

To establish well-posedness for the Caputo fractional SEIHRD model, we work on the Banach space

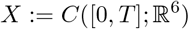

endowed with the supremum norm ∥*x*∥_∞_ = sup_*t*∈ [0,*T*]_ ∥*x*(*t*) ∥. The formulation of Caputo allows you to convert your differential system into an equivalent Volterra-type integral equation. This is the basis of the standard analysis of Caputo-type problems and is what allows you to use fixed-point methods in a straightforward and quantitative way [14,15,16]. Let *x*(*t*) = *S*(*t*), *E*(*t*), *I*(*t*), *H*(*t*), *R*(*t*), *D*(*t*)) ^⊤^. Denote *F* (*t, x*) be the vector field on the right-hand side of the equation from the model (including the nonlinear incidence through *λ*(*t*))). Under the regularity conditions that have been used in the previous sections (i.e., continuity in *t* and local Lipschitz continuity with respect to *x* in biologically invariant domain, Ω), the following initial value problem is defined:

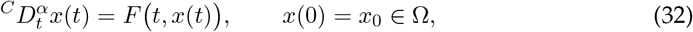

is equivalent to the integral equation

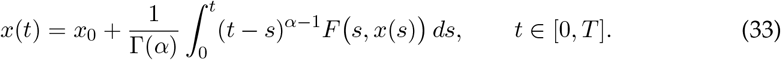

This representation motivates the integral operator *T* : *X* → *X* defined by

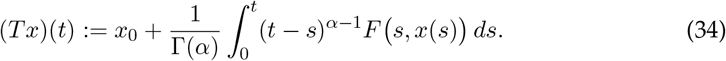

A solution to the fractional system therefore is a fixed point, *T*, and our job is to check that the conditions of the fixed point theorem hold on a properly closed subset of *X* [16].

### 5.2 Lipschitz condition and contraction principle

Assume that *F* is Lipschitz continuous in *x* on Ω uniformly in *t* ∈ [0, *T*]; that is, there exists *L* > 0 such that

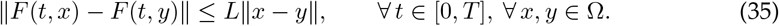

Then, for any *x, y* ∈ *X* with values in Ω, the operator *T* defined in (34) satisfies

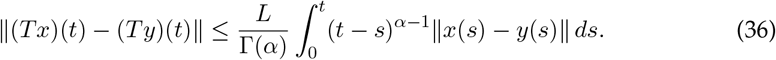

Taking the supremum over *t* ∈ [0, *T*] and using 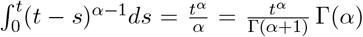 yields the uniform estimate

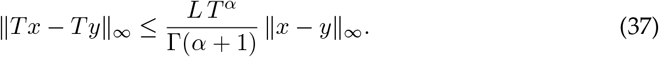

Consequently, a clean and verifiable sufficient condition for *T* to be a contraction on an appropriate closed subset of *X* is

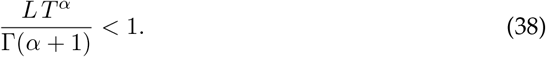

This inequality has an important application in that it shows how the admissible time horizon *T* is determined by the memory order *α* and Lipschitz modulus *L*. A great deal of the time, this inequality is enforced, for example, by either correctly solving the solution on a sufficiently short interval and extending it incrementally to reduce the effect of the error or by establishing refined one-sided bounds that decrease the effective Lipschitz constant on Ω, respectively [16].

### 5.3 Existence–uniqueness theorem and a compactness alternative

Under the above assumptions of the continuity of *F* with respect to *t*, Lipschitz continuity of F with respect to *x* on the set Ω, and the contraction condition *LT* ^*α*^*/*Γ(*α* + 1) < 1, Banach’s fixed point theorem ensures the existence of the unique fixed point of the operator *T* in the appropriate closed set of *X*. Hence, the Caputo fractional SEIHRD model has a unique solution *x*(·) ∈ *C*([0, *T*]; Ω) on the time interval [0, *T*]. The solution is continuously dependent on the initial data, and this provides the foundation for the simulation and robustness analysis, as the forward map *x*_0_ ↦ *x*(·) is well-defined and stable under perturbations [16]. However, in the case where a global contraction on the whole interval [0, *T*] cannot be guaranteed, for example, when large parameter ranges or large time horizons are desired, one can use compactness-based fixed point arguments after a priori bounds have been derived. A basic approach is to prove that *T* maps a bounded, closed, and convex set to a relatively compact set and that *T* is continuous. Then Schauder’s theorem guarantees the existence, although not uniqueness, of solutions. This alternative approach is well-known in the study of nonlinear integral equations and is often used in the study of fractional models where the Lipschitz constants become too large to ensure a contraction mapping globally [15,16]. In any case, the operational criterion for the assumptions will be that they can be immediately checked from the epidemiological vector field, and this will often involve combining the results on positivity/invariance (Section 4) with explicit bounds on the nonlinear incidence in Ω. These “auditable” hypotheses are not superficial because they guarantee that the mathematical results actually support the computational and epidemiological interpretations and do not simply amount to a form of formalism [16].

## 6 Equilibria and Threshold Dynamics

### 6.1 Disease-free equilibrium (DFE) and the role of spillover

Threshold analysis for compartmental epidemic models usually revolves around the diseasefree equilibrium (DFE), which refers to a state where there is no ongoing transmission in the human population. For models that incorporate external inputs, the notion of “disease-free” is heavily dependent on whether external inputs are assumed to be zero. In our model, the effect of spillover is represented by the non-negative input *η*(*t*), which generates new exposures through the *S* → *E* pathway. The model has a natural decomposition into two regimes that can be considered analytically distinct. First, when spillover is absent, i.e., *η*(*t*) ≡ 0, the classical DFE is well-defined as the equilibrium with *E*^∗^ = *I*^∗^ = *H*^∗^ = 0, *R*^∗^ = 0, *D*^∗^ = *D*_0_ (or *D*^∗^ = 0 if deaths are initialized at zero), and *S*^∗^ = *N*_0_ (or more generally *S*^∗^ = *N*^∗^ consistent with the conserved bookkeeping). In this setting, the DFE actually represents a transmission-free state for the human-to-human subsystem, so that the associated reproduction threshold *R*_0_ measures the intrinsic transmissibility of the virus in the population in the absence of any importations [18,19]. Secondly, if the spillover effect persists (*η*(*t*) ≢ 0), a strictly disease-free steady state is not generally meaningful in the literal sense because the system is constantly being forced away from the equilibrium *E* = *I* = *H* = 0. In such cases, the appropriate conceptual framework is the imported infections perspective, in which one can clearly distinguish the endogenous level of amplification capacity, i.e., the endogenous level of transmission potential, as measured by the reproductive metric with *η* ≡ 0, from the exogenous forces that drive the dynamics and ensure a non-zero level of incidence even in the absence of endogenous transmission potential [18,19]. In practice, we thus define the DFE and its stability for the autonomous baseline, i.e., for the parameter value *η* ≡ 0, and then interpret the time-dependent function *η*(*t*) as an external driver, whose epidemiological effects are evaluated in terms of scenarios rather than in terms of equilibrium eradication statements.

### 6.2 Basic reproduction number *R*_0_ via the next-generation matrix

The basic reproduction number is calculated by the so-called Next Generation Matrix (NGM) method for the infected subsystem around the DFE with *η*(*t*) ≡ 0. This is the standard way for compartmental models and offers an easy-to-interpret decomposition into “new infections” and “transfers” among infected states [18,19]. In our model, the infected-state vector can be taken as (*E, I, H*) since these classes mediate the creation and propagation of infection. Linearizing the infected subsystem around the DFE yields a decomposition of the form

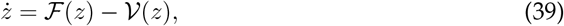

where ℱ collects the appearance of *new infections* and 𝒱 collects all other transitions (progression, hospitalization, recovery, mortality). The NGM is then *K* = *FV* ^−1^, where *F* and *V* are the Jacobians of ℱ and 𝒱 evaluated at the DFE, and the basic reproduction number is

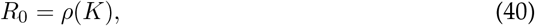

the spectral radius of *K* [18,19]. An important methodological aspect of the current study is that the obtained R0 is written down explicitly in terms of the parameters of the epidemic process (including *β* and *κ* describing community and residual hospital transmission, and *ρ* controlling the routing of infectious individuals into isolation). While this explicit dependence may be considered merely a matter of closed-form appearance, it is actually crucial to later global sensitivity analysis, as this explicit dependence enables a clear quantification and ranking of the contribution of each parameter to the invasion threshold [7, 11]. In particular, isolating the role of hospital-related transmission through *κ* allows for the distinction between “community-driven” and “care-setting” amplification, which is meaningful in the context of Nipah outbreaks where infection control is a decisive intervention lever [2,13].

### 6.3 Local stability of the DFE in fractional-order systems

Stability of local equilibria in Caputo fractional systems must be established through criteria different than those used to evaluate integer-order systems. In an example of a fractional-order autonomous system, with *α* ∈ (0, 1)) viewed at an equilibrium point will provide a matrix *J* used to identify the eigenvalues to be evaluated for determining stability according to the fractional equivalent of exponential decay. The DFE is locally asymptotically stable when all eigenvalues *λ* of the Jacobian satisfy

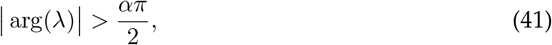

which is the standard sector condition for Caputo dynamics and reduces to the familiar “negative real part” criterion when *α* = 1 [14,15,16]. This condition in epidemic models is typically guaranteed to hold true by demonstrating that the linearization of the infected sub-system is stable when the basic reproductive number *R*_0_ < 1, thereby preventing small initial perturbations from resulting in the invasion of infection when *R*_0_ < 1. From the construction of NGM, the classical threshold dichotomy exists in a manner that is consistent with fractional dynamics: For *R*_0_ < 1, we have local asymptotic stability of the DFE (i.e., cannot have sustained growth of very small seeds of infection) and *R*_0_ > 1 means that the DFE is unstable (i.e., can be invaded) [18,19]. Spillover has different meanings based on context, which is significant. That is why the local stability of DFE under continuous introduction of cases (the condition for local stability is *η* ≡ 0) does not imply “eradication” under presence of new cases (i.e. when *η* ≢ 0); it indicates that cases imported in this manner will not cause self-sustained amplification (due to the force) beyond the force’s direct effect; and the trajectories will stay bounded in all instances of a positive input, which is consistent with the concepts of robustness and Ulam hyers stability defined below.

### 6.4 A route toward global stability (or a principled substitute)

Global stability proof is far more complicated for nonlinear fractional epidemic models than local analysis; you need to use some form of Lyapunov argument that is modified to include non-integer derivatives and comparison principles. One way of providing strong results is constructing a fractional Lyapunov functional *V* such that along solutions its Caputo derivative shows signs which guarantee that trajectories will converge towards the DFE (with *η* ≡ 0 and *R*_0_ < 1). The use of such techniques has been found in the larger body of literature on fractional dynamical systems, and their success in epidemic applications is limited to those with appropriate incidence structure and invariant region that result in sharp inequalities [14,15,16]. When it is not possible to obtain a global theorem with full rigor or to create any results from general ranges given, an appropriate alternative is simply to state specifically what has been shown without restriction; these are global boundedness/invariance (Section 4) and local stability (this section), followed by demonstrating numerically whether there are trajectories with global attraction among the parameter spaces of interest. The strategy of using “global boundedness + local stability + numerical verification” should not be considered as a replacement to theory but can be used as an acceptable and clear method of transitioning when the full construction of a Lyapunov function is not possible due to needing to keep the model as close to actual epidemiologic conditions and with usable parameters [15,16]. Our goal has been to move towards a comprehensive analysis of the subject matter. However, we must do so without compromising the assumptions which can be audited by using the model’s epidemiological vector field as reference.

## 7 Ulam–Hyers and Generalized Ulam–Hyers Stability

### 7.1 Approximate solutions and an explicit error model

For a Caputo fractional epidemic system of order *α* ∈ (0, 1),

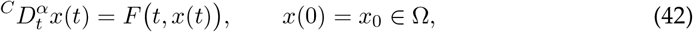

Ulam-Hyers Stability applies the requirements of robustness to the processes used in modelling: a simulator’s or even an imperfect dataset’s output trajectory will generally not be an exact solution; however, it should have a uniformly small error with respect to an accurate trajectory provided that the error in the model governing laws known as “defect” is also uniformly small.

Accordingly, we call *y* ∈ *C*([0, *T*]; Ω) an *ε*-approximate solution if its residual satisfies

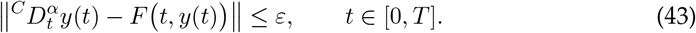

*ε* can represent (i) structural mismatch (missing heterogeneity, neglected pathways), (ii) discretization/rounding inaccuracies, and (iii) data incompatibilities caused by estimating model parameters from sparse or delayed observations. The original stability issue arises from the resolution of Ulam’s Problem by Hyers and its subsequent extensions in the direction of Hyers-Ulam-Rassias [4,10]. The fractional variants of these results utilize the equivalency between the Caputo and the Volterra integral as well as fractional Grownwall-type control inequalities, converting small residuals directly into small trajectory deviations with an explicit constant [8,14,15,16,17].

### 7.2 Main Ulam–Hyers stability theorem (fractional Caputo setting)

Assume:

1. **(Well-posedness setting)** Ω ⊂ ℝ^*m*^ is a positively invariant region for the model, and *F* : [0, *T*] × Ω → ℝ^*m*^ is continuous in *t*.
2. **(Lipschitz condition in the state)** There exists *L* > 0 such that 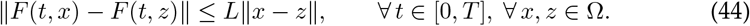
3. **(Approximate solution)** *y* ∈ *C*([0, *T*]; Ω) satisfies

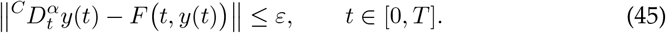

Let *x* be the exact solution of the IVP with the same initial data *x*(0) = *y*(0) = *x*_0_. Then one obtains an explicit uniform bound

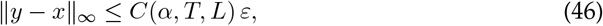

where a standard admissible choice of robustness constant is

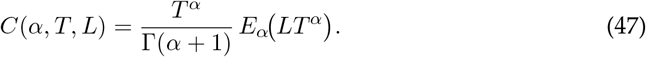

#### Why this constant appears (mechanism)

Writing both dynamics in integral form, the defect bound implies an inequality of the type

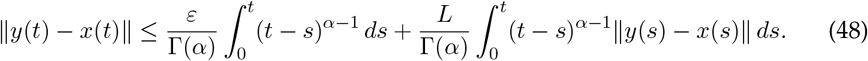

The initial integral can be represented as a product of *t*^*α*^*/*Γ(*α* + 1). The second term is bounded using the Fractional Gronwall inequality, producing a Mittag-Leffler function *E*_*α*_(*Lt*^*α*^) giving rise to the uniform bounds between equations (46)-(47) [14,15,16,17]. Therefore, *Caputo integral representation + Lipschitz continuity + fractional Grönwall* ⇒ *quantitative robustness*.

### 7.3 Generalized Ulam–Hyers stability

Generalized Ulam–Hyers stability replaces the linear control *ε* → *Cε* by a broader error-class function *ϕ*, reflecting that modeling and data errors may scale nonlinearly.

Assume the same hypotheses as in Section 7.2, but replace the residual bound by

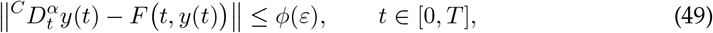

where *ϕ* : [0, ∞) → [0, ∞) is nondecreasing and *ϕ*(0) = 0. Then

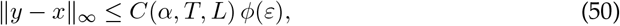

with the same *C*(*α, T, L*) as above. The original conceptualization of “stability” has been preserved in a generalized form (uniformly bounded defects enforce uniformly bounded deviations) whereas; in terms of introducing residual aggregates of heterogeneous sources of uncertainty (measurement noise+discretization+parameter perturbations), it is far more representative of what we would have expected [8,10,14,15,16,17].

### 7.4 How to read the bounds in practice (link to sensitivity and intervention levers)

The constant *C*(*α, T, L*) is not a decorative artifact: it identifies which modeling regimes are intrinsically *fragile*.

- **Reliance upon** *L* **(epidemiological rigidity):** Higher Lipschitz modulus values correspond to an increased amplification of any deviations from equilibrium by the vector field, so the deterioration of the Ulam-Hyers bounds occurs. In compartmental models, this indicates an increase in *L* as the transitional rate increases (i.e., *β* increases), as the strength of the nonlinear incidence response increases, and as the radius of the invariant region increases (i.e., the range of states in which solutions can lie increases).
- **Dependence on** *T* **(forecast horizon):** Robustness is weaker as the forecast horizon becomes longer; even for small values of *ε*, the total effect of the memory over the interval [0, *T*] can be represented as *T* ^*α*^*E*_*α*_(*LT* ^*α*^).
- **Dependence on** *α* **(memory regime)**. When *α* is altered, the type of propagation is impacted: the Mittag–Leffler envelope can be interpreted with respect to the “slow” algebraic-type (fractional) relaxation as *α* ↑ 1 or classical exponential behaviour. In practice, robustness will be a function of both the memory and the regime that has been selected as opposed to “individually by parameter”-that is, [14,15,16,17].

The conceptual link from global sensitivity to global sensitivity analysis includes two aspects. One is sensitivity ranking to identify which parameters drive desired outcomes. The other is Ulam–Hyers stability, which looks at which parameter regimes will increase the magnitude of any error in the model as evidenced by trajectories deviating from model predictions. If an intervening parameter (for example, isolation effectiveness or hospitalization-related infection spread) lowers the effective Lipschitz amplification in the infectious disease subsystem dynamics, both (i) dynamical controllability and (ii) margins of robustness increase. This provides a rational basis for prioritizing intervention parameters in the presence of uncertainty [7,11].

## 8 Sensitivity, Identifiability, and Uncertainty Quantification

### 8.1 Local sensitivity (analytic)

In order to understand the mechanisms through which threshold behavior occurs, we will start by determining how sensitive the basic reproduction number *R*_0_ is to a local (derivative-based) change in the parameter *p*. Since *R*_0_ is dimensionless, the standard and interpretable choice is the normalized sensitivity index

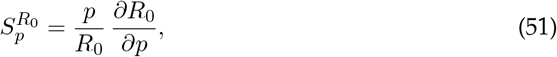

which measures the relative change in *R*_0_ induced by a relative perturbation of *p*. If the *R*_0_ value is present either as exact (or semi-exact) values using methods for generating next generation matrices will allow for analytical computation of the indices based on stable symbolic/automatic differentiation yielding a quick computed ranking list of “critical parameters” in proximity to the baseline regime [18,19]. Calculating a locality index has multiple benefits but one key benefit of calculating the locality index is that it provides a guide to whether *R*_0_ is driven by community-based transmission, care-based amplification, or care-based isolation related parameters, and thus, provides a way to help establish plausibility for identifiability and develop global uncertainty studies.

### 8.2 Global sensitivity (PRCC or Sobol)

Neighborhood-based sensitivity is termed local; epidemic discovery & decision support typically need robust results across many defensible parameter ranges. Therefore, we apply global sensitivity analysis using each of the uncertain parameters considered herein to create a random variable for that parameter containing all values from a biologically and clinically acceptable interval defined in section 3.3. The implementation of Latin Hypercube Sampling (LHS) is a method of sampling from the parameter space in a manner that allows stratification of inputs that have monotonic (or nearly monotonic) input and output relationships [7,11]. The method allows us to compute Partial Rank Correlation Coefficients (PRCC), which are coefficients that represent the degree and direction of the relationship between each input and output, controlling for the other input values [7]. Similarly, for interactions that dominate outcomes and for outcomes with nonmonotonic relationships, we are using an approach based on variance decomposition to calculate Sobol indices that split empirical output variance into first-order and higher-order contributions [11]. We conduct our analysis on outbreak-related outputs that account for at least two different aspects of each output, specifically thresholds and burdens; R0 at its autonomous baseline where *η* ≡ 0, Imax at peak intensity, tmax at peak time and cumulative mortality *D*(*T*). The results have been presented graphically to support visual representation of this by means of compact summary visualizations (e.g., *PRCC* heatmaps generated from outputs or Sobol bar plots), thereby providing a direct comparison of (i) the parameters that mostly affect the relative ability to invade and (ii) the parameters that heavily influence maximum loads and severe outcomes - two aspects which are inconsistent, due to the nonlinear, memory-based nature of dynamic systems.

### 8.3 Identifiability as a methodological checkpoint

As a rule, sensitivity and identifiability cannot be equated; even if a variable is very important, if data do not exist to adequately identify the variable it will nonetheless remain weakly important. We regard identifiability as a methodologic checkpoint to show what can be learned from calibration, and also what remains uncertain. To determine if a parameter vector is uniquely identifiable - that is recoverable in the theoretical absence of noise - from the model outputs with the selected observation operator is a structural issue related to the identifiability of the parameter vector. Compartmental epidemic models are notoriously difficult to work with structurally, even without a complete symbolic treatment. The following are typical structural risks for these models: 1) confounding of parameters (e.g., product and ratio based); 2) indistinguishable progression and removal pathways; and 3) non-uniqueness as a result of partial observability of the latent compartments. Practical tests of identifiability are performed with respect to realistic data noise and the potential limited sample sizes. If the data available allow, we recommend using profile likelihood diagnostics to identify flat directions and non-unique confidence intervals, and/or bootstrap based uncertainty assessment for fitted parameter estimates, particularly when calibration is being performed against short time periods of incidence and/or aggregated incidence series. The checkpoint verifies that sensitivity rankings are interpreted correctly. A parameter can be flagged as a “dominant” parameter; however, if it does not have a well-defined identity, it should be prioritized on the data enrichment matrix (e.g. by hospitalization time series, serology, or contact structure proxy) as a primary target for data enrichment. In other words, it should not be viewed as a credible lever based solely on limited incidence evidence in regard to parameter classification.

### 8.4 Uncertainty propagation and credible outcome bands

In the end, we have transferred uncertainty associated with the parameters in the model to the associated uncertainty with the outcomes of that model to provide decision envelope estimates instead of a single trajectory. To do this we have simulated the model outputs using the same parameter sample sets as used in the global sensitivity study and report the uncertainty bands around those simulated values for the key quantities of interest from the model. Since epidemic outcomes can be very variable, we provide a summary of their growth over time as well as their overall performance with robust statistics (for example, median values, plus or minus the central 95% credible intervals). We consider this envelope to reflect some of the operational uncertainties caused by parameter uncertainty; spillover scenarios for a given outcome, denoted as *η*(*t*); and memory regimes for this outcome, denoted as *α*. This step closes the loop. Ulam-Hyers bounds provide assurance of robustness against modeling errors at the dynamical level, while uncertainty propagation provides an estimate of how uncertainty in the parameters will result in uncertainty in the public health endpoint. This allows a transparent comparison of scenarios and allows for prioritization of interventions in the absence of complete information [7,11].

## 9 Numerical Scheme and Verification

### 9.1 Choice of the numerical method

In order to efficiently simulate the Caputo SEIHRD dynamics of a fractional order on a finite time period [0, *T*], we plan to use a predictor-corrector Adams scheme specifically designed for the Caputo fractional order. The key idea is to represent the initial value problem as a Volterra integral,

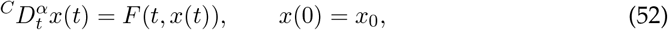

which is equivalent (under the regularity assumptions already stated in the preliminaries) to

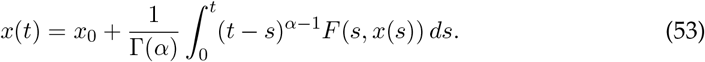

Not only is the integral form easy to manipulate mathematically, but it naturally lends itself to numerical discretization because it clearly identifies the memory kernel and immediately converts the evolution problem into a quadrature problem that is dependent upon history. The PECE process is a predictable way to bring about an algorithm that allows for both the predictive and corrective possibilities of predicting diseases through their associated vectors. This is beneficial as it keeps the vectors from being altered by any type of smoothing operations. In numerical analysis of Caputo problems, this is a standard technique and is often used to solve problems using classic numerical techniques [16, 20]. In particular, let *t*_*n*_ = *nh* for *n* = 0, 1, …, *N*, *h* = *T/N*. The aim is to find *x*_*n*_ ≈ *x*(*t*_*n*_) using a two-step iterative method: (i) first find a predictor for *x*_*n*_ using an Adams–Bashforth type product-integration rule and (ii) then determine the corrected value of *x*_*n*_ through an Adams–Moulton type refinement. The approach is very appealing for our problem since it (a) respects the integral structure imposed by the Caputo operator, (b) treats nonlinear incidence functions without linearization, and (c) provides a straightforward approach to the verification of mesh refinement [16,20]. There are higher-order versions of the product integration/extrapolation methods, as well as other quadrature methods, but we will concentrate on the robust basic ABM predictor-corrector method in the main text, reserving comparisons of this type (including higher-order interpolation formulas) for the appendices as necessary [16,20,21,22].

### 9.2 Convergence considerations and practical error control

As a result of this history accumulation in fractional dynamics, the reliability of the numerical solution depends on the control of the local quadrature error as well as the memory accumulation. This can be achieved in a structured way by the predictor-corrector approach. This approach uses a predictor step for a provisional state 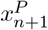, followed by a corrector step. For the Caputo integral equation, the PECE scheme can be written in the standard weight form

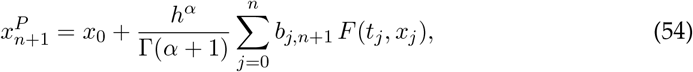

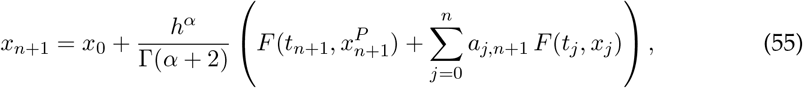

where the weights {*b*_*j,n*+1_} and {*a*_*j,n*+1_} are explicit and depend only on *α* and the step index (hence are precomputable). The “Adams-type” structure of PECE will be used as a reference schema for this study of fractional initial-value problems, as it is commonly utilized in research [16,20]. Error control and verification will be accomplished through multiple associate mechanisms to improve the efficiency of each technique in reducing computation time and errors [16,20].

#### 1. Mesh refinement (step-halving) test

The solver should be executed for [0, *T*] utilizing step sizes *h* and *h/*2 then compare key outputs (for example, *R*_0_ related quantities, *Imax, tmax, D*(*T*)) and where necessary compare entire trajectories using the sup-norm. The constancy of the contraction of the difference is the best benchmark of verification for fractional simulations because it eliminates the effect of memory, which will also eliminate any artifacts introduced by discretisation [16,21].

#### 2. Defect (residual) monitoring

Since Ulam–Hyers analysis is fundamentally a “small defect ⇒ small deviation” principle, it is natural to compute a numerical defect indicator at discrete times, e.g.

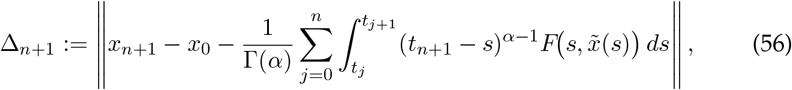

with 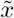 taken as the piecewise approximation used by the scheme. Practically, max_*n*_ Δ_*n*_ which represents the maximum error of the quadrature structure and can be used to assess whether the numerical defect is sufficiently small (i.e., the numerical errors were present in all numerical results) for subsequent judgments regarding numerical stability/sensitivity to be trustworthy.

#### 3. Positivity and invariant-set preservation checks

Biological feasibility of the compartment solutions must always be ensured by validation of each time step such that *S*_*n*_, *E*_*n*_, *I*_*n*_, *H*_*n*_, *R*_*n*_, *D*_*n*_ *≥* 0.; if small negative value(s) are observed (most often resulting from coarse time steps in a region of high transience), it is more effective to decrease the size of the time step (*h*) rather than to use excesive projection techniques as projection can negatively impact convergence rate and unitary conceptualization. We will also verify that the bookkeeping relationship defined analytically (i.e. conservation of *N*(*t*) + *D*(*t*) in a closed accounting framework) continues to be valid at each discrete time point as a key diagnostic tool for identifying any implementation error.

##### Computational cost note

Directly implementing memory sums maintains an *O*(*N* ^2^) scaling because they are historically dependent. For moderate horizons with standard epidemiological calibration grids, that is acceptable. For lengthy horizons or large ensembles (e.g. in global sensitivity), efficient implementations of memory sums must be considered (e.g., re-use of weights, vectorization, and when appropriate, fast convolution methods) with the same numerical scheme [21].

### 9.3 Reproducibility package and verification protocol

To make the numerical claims auditable and reusable, we provide a reproducibility bundle organized around three layers:

#### 1. Parameter and scenario specification (machine-readable)

All parameters (including *η*(*t*), intervention levers such as *κ* and routing parameters, and fractional order *α*) can be found in an individual configuration file (YAML/JSON). Each rundependably stores logged parameters: used parameter vector, used initial condition, *T*, used *h*, and spillover choice.

#### 2. Versioning and deterministic execution

This version of code is backed by a version-control system, and a commit hash (or release ID) is appended to each figure or table that is shown in the manuscript. Monte Carlo / LHS ensembles record the random seed, the sampler settings and the complete list of sampled parameter sets, meaning that all sensitivity plots can be re-generated with the same results (byte-for-byte).

#### 3. Verification checklist (reported in the manuscript or supplement)

The following items will be reported: (i) Positive satisfaction rates and worst-case negative satisfaction rates (should be essentially zero when discretization has been verified); (ii) discrete conservation/identity errors in the values of *N*(*t*)+*D*(*t*); (iii) differences due to mesh refinement for outputs of primary interest; and (iv) runtime and cost metrics for ensemble studies. This checklist describes standard reproducible work practice in computational research; this is needed to separate model uncertainty from numerical uncertainty and to avoid excessive interpretation of discretization artifacts [23,24].

## 10 Numerical Experiments

In this manuscript each experiment was conducted to provide definitive evidence regarding the validity of a mechanistic hypothesis (i.e., memory, spillover forcing, or intervention leverage) by providing clear comparisons between observed experimental outcomes and the corresponding underlying mathematical model. The numerical simulation methodology followed a Caputo tailoring of the predictor-corrector integration method focusing on verification through refined measurements, defect detection, and identification of model-experimental differences due to discrete artifact [14,16,21]. All experimental results included a comparable set of outbreak relevant summary statistics including threshold, maximum size, maximum timing, cumulative mortality, and attack rate estimates [18,25,26] for comparison among the various regimes.

### 10.1 Baseline dynamics (nominal parameter set)

To begin with, it is essential to define a nominal parameter vector that lies within biologically rational parameters as previously delineated and therefore generate a simulated (full state) trajectory [0, *T*] (state variables) of (*S, E, I*) by (*S, E, I, H, R, D*) over the course of [0, *T*] would have two purposes: First, to permit interpretation of the results of all future perturbations using reference data; and secondly, to provide evidence that when using a numerical solution, the conditions necessary to maintain an admissible region (i.e. no negative numbers) and a closed bookkeeping identity (i.e. *N*(*t*) + *D*(*t*)) are within the limit of numeric resolution tolerances [14,16,21]. Thus the base case values can be thought of as a calibration of empirical value (i.e. time intervals for latent build-up, symptomatic rise in cases, level of hospitalizations and accumulation of severe outcomes) as well as providing assurance that the numerical solution produces results consistent with the qualitative compartment(s) of the model [18,25,26].

**Figure 1.**
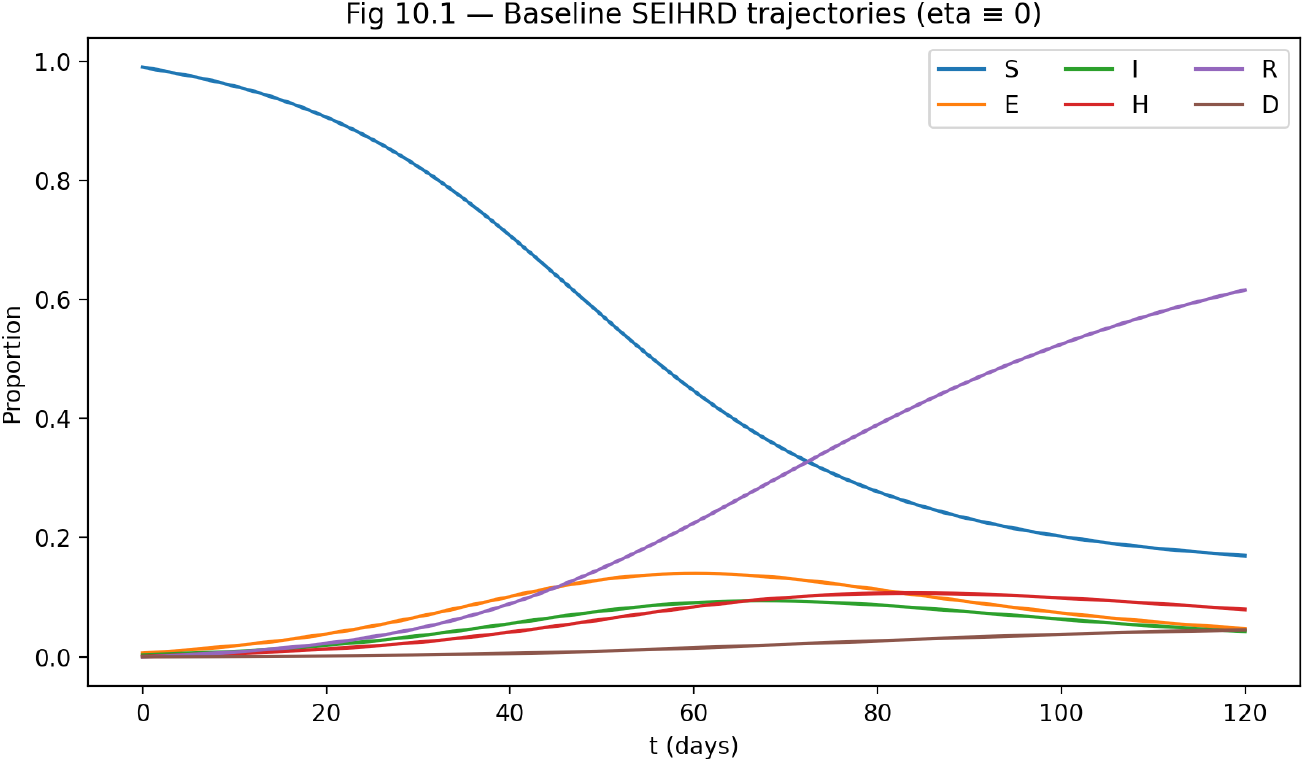
This figure establishes the *reference outbreak trajectory* of an outbreak without outside influence (no external force of infection) as described by a nominal parameter set without exogenous spillover forcing (*η* = 0) of combined parameters. The susceptible compartment, denoted *S*(*t*), declines smoothly and monotonically over time. This is indicative of an on-going transmission process that will eventually be interrupted by the exhaustion of the susceptible population. The exposed and infected compartments, denoted *E*(*t*) and *I*(*t*) respectively, will progress through the latent and infectious phases in accordance with the model’s anticipated structure of incubation; as such, *E*(*t*) will peak earlier than *I*(*t*) and at lower amplitude than *I*(*t*). While the total number of hospitilized patients *H*(*t*) in the community increase at a slower rate and peak later than community transmission, due to the clinical lag time in the burden of hospitalizational illness. As there are an increasing number of individuals with recovered from illness *R*(*t*) that will eventually dominate the overall population; however, the cumulative dead *D*(*t*) will be growing at a constant rate and be bounded for the entirety of the simulation period. It is significant that the order of the peaks is maintained chronologically (i.e., *E* → *I* → *H*—is preserved). Thus, we can conclude that the solver produced a method that preserved the intent of the original model through qualitative compartmental reasoning and did not create numbers artificially. In addition to providing a descriptive dynamic, the baseline trajectory will also be the reference calibration point that measures and provides a benchmark to all perturbations (fractional order, spillover force, and intervention) in order to assure that differences observed can be relied on to be due to the controlled mechanisms of the model and not due to some inherent instability of the baseline.

**Figure 2.**
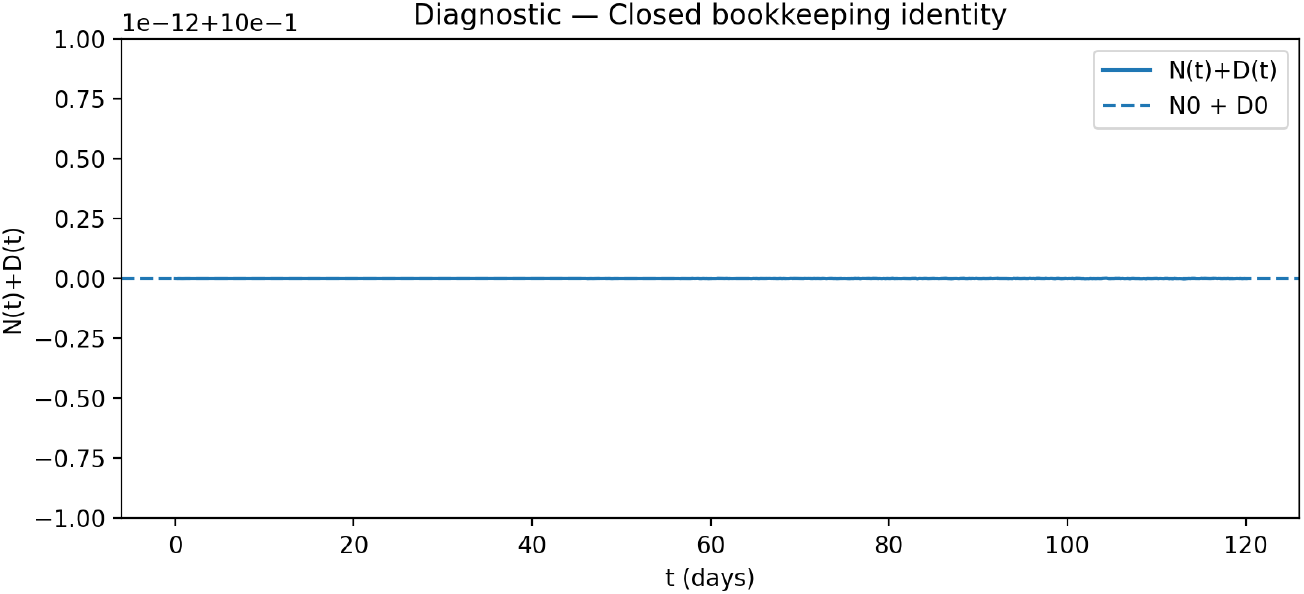
This figure provides a *numerical verification* of the closed bookkeeping identity, tracking the quantity *N*(*t*) + *D*(*t*) over time. The trajectory remains effectively constant (up to machine precision) across the entire integration window, indicating that population mass is conserved modulo disease-induced mortality, exactly as prescribed by the analytical model structure. This result is not merely cosmetic: in fractional-order systems, long-memory accumulation can amplify small quadrature errors into spurious drift. The absence of such drift here confirms that the Caputo-tailored predictor–corrector scheme respects the invariant accounting structure at the discrete level. Consequently, the baseline dynamics in Fig. 10.1 can be interpreted as *physically and epidemiologically admissible*, providing a reliable foundation for sensitivity analysis, spillover scenarios, and robustness testing in the subsequent sections.

### 10.2 Effect of the fractional order *α* (memory regime)

To determine how much memory affects performance, we will change the value of *α* from a range of (0, 1), using only data collected during the same time period (while keeping all other variables constant). After changing the value of *α*, we will compare the result values created by using only data collected with a different value of *α*. The testable hypothesis is that lower values of *α* will lead to the restructuring of epidemic growth rates because the Caputo memory kernel will change the way that previous values affect current growth rate values. This effect generally will show up as changes in the distribution of time spent in the growth and relaxation phases; i.e., time rescaling will be different for each (growth or relaxation) phase, rather than uniformly [14,16]. We show the movement of both peak intensity *I*_max_ and peak time *t*_max_ as they vary according to *α*; these movements can be interpreted as evidence of historical persistence. Changes in the size of *α* affect the degree of historical influence. Lower *α* causes greater historical influence, which may alter the timing and/or shape of the peak Imax, and the shape of the burden profile may also change as a result of the lower *α*, even though the instantaneous vector field is the same ([14,16]). Note that all comparisons are made using exactly the same spillover and intervention conditions, so that any difference observed is caused only by differences in memory.

**Figure 3.**
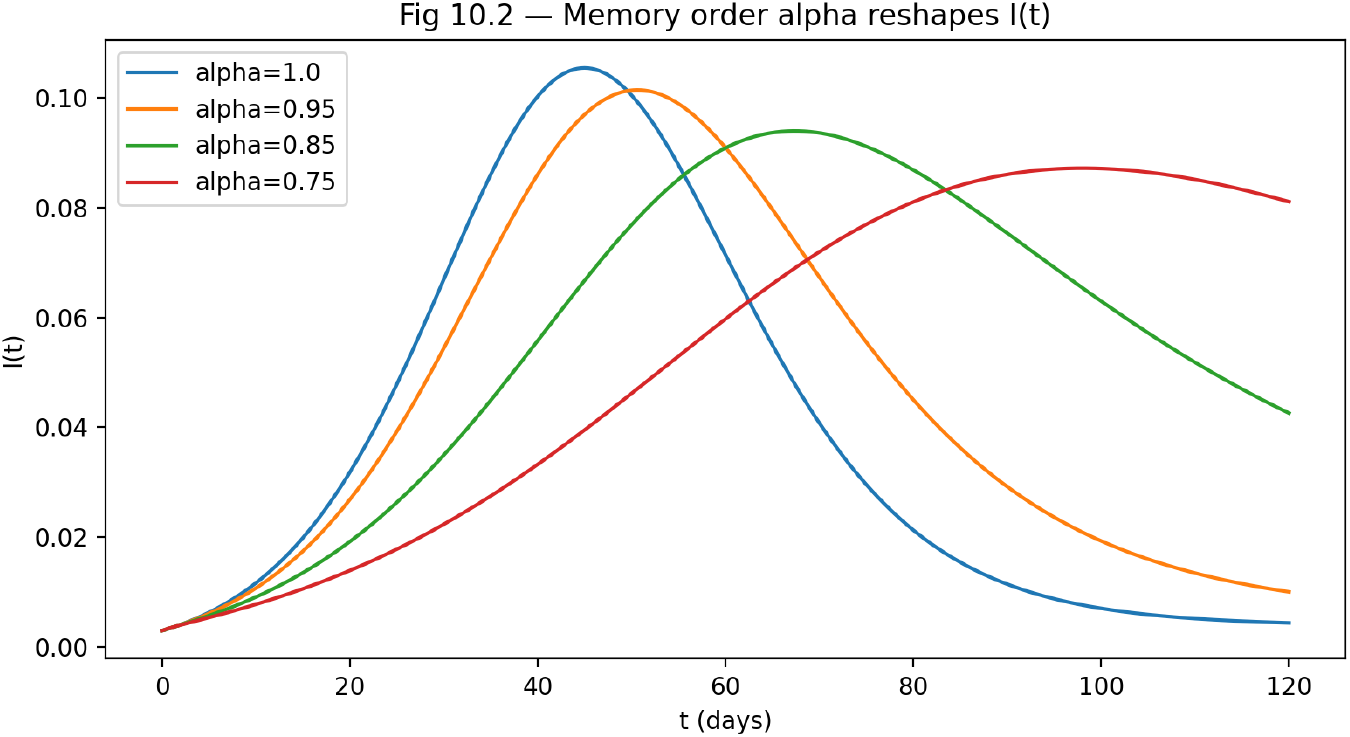
The Impact of Fractional Order *α* on Infection Dynamics. When using a different fractional order *α*, the temporal shape of the infectious class *I*(*t*) will be affected. Lower values of *α* have a greater effect from historical data, causing peaks to occur later, outbreak profiles to be wider and longer lasting, and by prolonging the existence of the infectious class. At the same time, the instantaneous vector field will not change. These findings demonstrate that memory will affect the outbreak kinetics through the Caputo kernel and not just through a rescaling in time, and will therefore support the interpretation of the effects of interventions in a manner that takes into account the impact of memory.

### 10.3 Spillover scenarios *η*(*t*): constant, impulsive, and seasonal forcing

Spillover is considered an externally generated input that creates exposure through the *S* → *E* mechanisms. Entry through the *S* → *E* channel allows researchers to evaluate amplification (endogenous) separate from external influences. Evaluating spillover impacts; we identify three archetypal forced driving factors. These include (i) an ongoing low level of background spillover, (ii) a series of impulse-like events with short durations and many exposures, and (iii.) a pattern of seasonal variations which can be associated with either environment-mediated contact or behaviourally-mediated contact. The baseline parameter set has been used to compare the two scenarios under uncertainty ranges, again to see if their robustness is affected. The primary question is not simply whether or not spillover will increase the burden of disease; that is simply a matter of speculation. The primary question, rather, is when and how the “spillover” of pathogens will shift the system from a transmission-dominated to forcing-dominated system, when “to eradicate” gives no real meaning to a statement about eradication, and when disease outcomes will better be interpreted through sustained imported pressure dynamics [18,25,26]. As such, we are able to assess differences between scenarios using threshold-based metrics calculated for the autonomous baseline (where *η ≢* 0), in addition to outcome envelopes for *η* ≠ 0, with a primary focus on how interpretable the results are in terms of public health planning (for example: sustained level of incidence after internally transmitted cases are less than what we observe).

**Figure 4.**
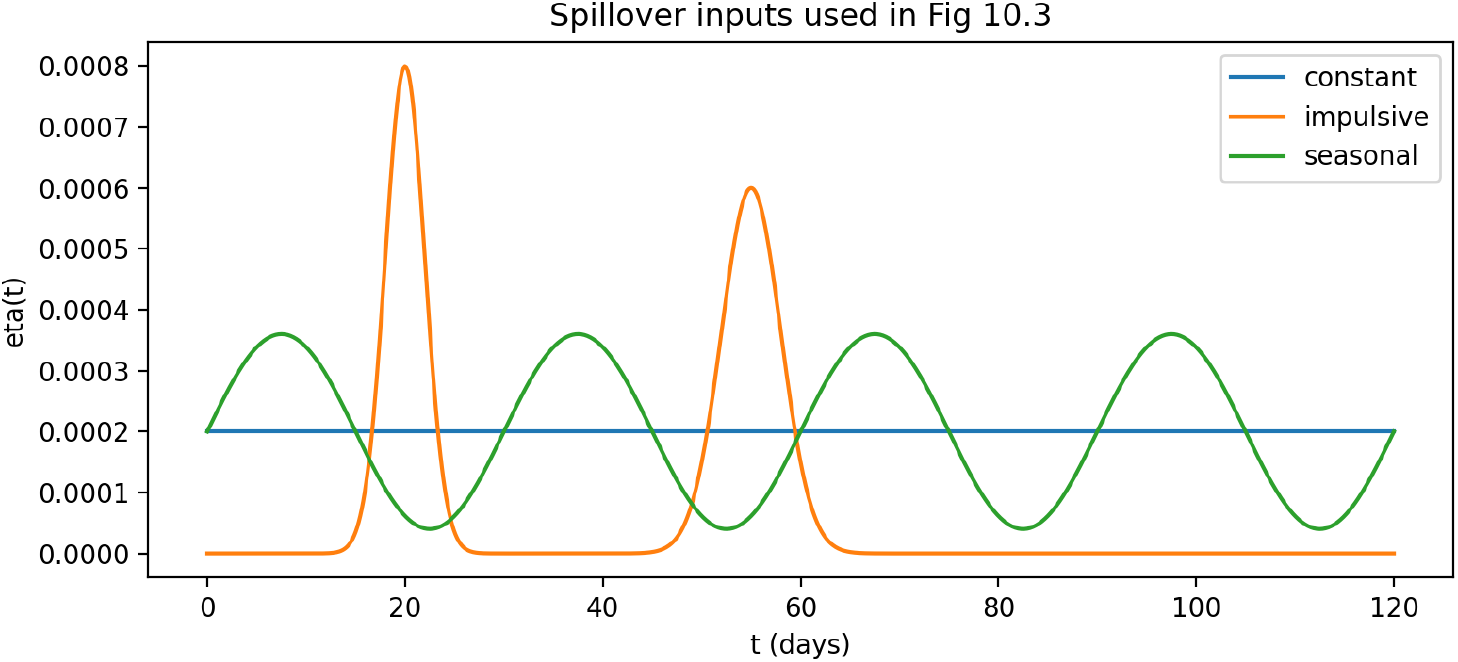
Spillover input functions *η*(*t*) used in Fig. 10.3: constant background forcing, impulsive exposure bursts, and seasonal modulation.

**Figure 5.**
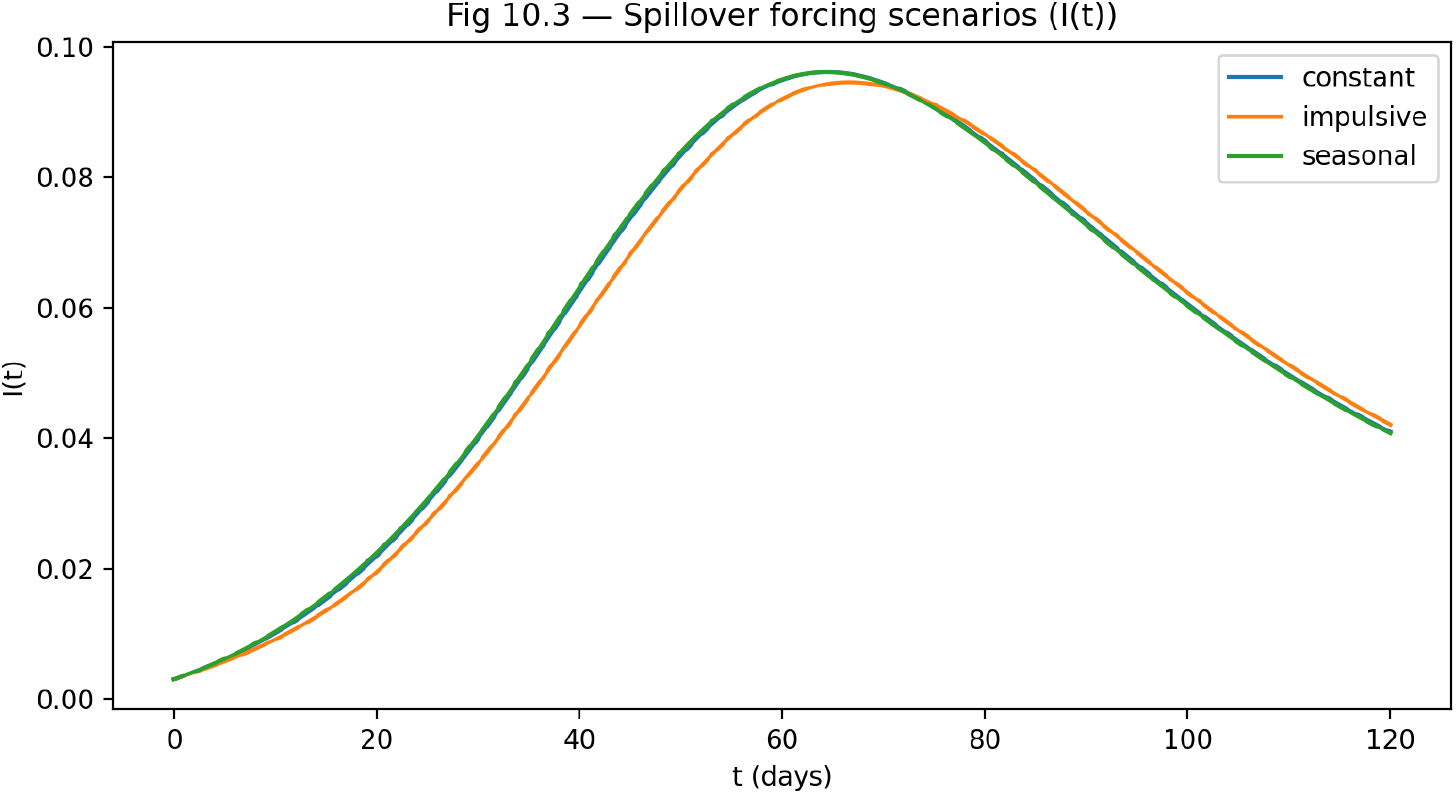
The effect of spillover forcing conditions on the number of infected individuals (*I*(*t*)) under identical baseline parameters. Differences between the scenarios arise due to timing and nature of external influences instead of differences in intrinsic transmission.

### 10.4 Numerical validation of Ulam–Hyers robustness (controlled defect injection)

This paper provides a uniquely valuable input in that the concept of robustness is substantiated by numerical testing under defined “stress” conditions. To accomplish this, we generate an *ε*-approximated trajectory *y* that results from imbedding a controlled defect in the governing equations, or equivalently, a bounded perturbation term (on the right side of the integral formulation). Accordingly, we compute the above definition of *ε*, through a residual/defect diagnostic that is consistent with the quadrature used for computation, and we also determine ∥*y* − *x*∥ _∞_, given the same initial conditions as the reference trajectory (See [14,16,21]). The method of determining if this hypothesis holds will be conducted through empirical testing. Specifically, we will be testing whether the deviations, ∥*y* − *x*∥ _∞_, scale in the predicted fashion as per the Hypothesis of Ulam & Hyers or how much worse they do than predicted. In addition, the testing will be for a given *C*(*α, T, L*)*ε* value with respect to how to relate to the memory and time values described in [14,16]. As a “numerical theorem check” we demonstrate this validation with a log-log graph of the deviation and the injected defect across multiple *α* values and horizons, as well as refinement tests to confirm that the robustness we see is the result of numerical formulation errors, not discretization artifacts [21]. This section thus establishes Ulam-Hyers stability as an operational measure for certifying reliability for simulation based conclusions.

**Figure 6.**
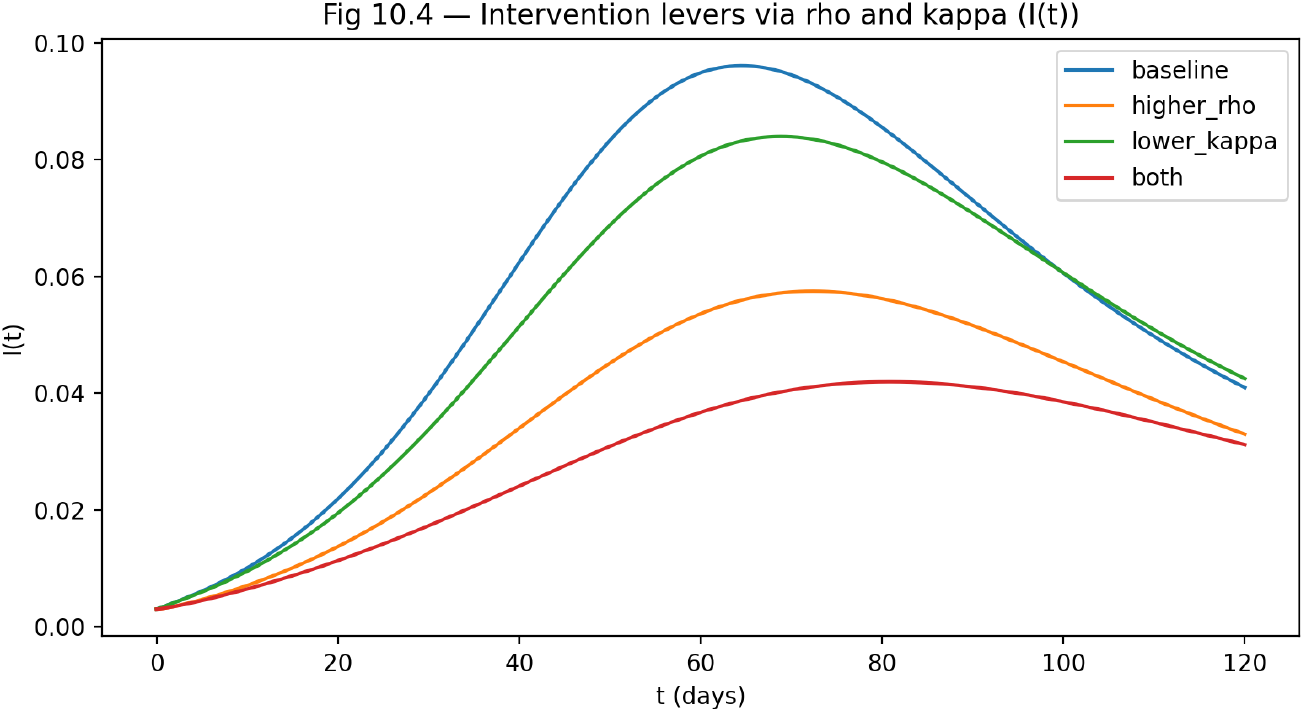
The figure presents the influence of two parameters (*ρ* and *κ*), that is, the rate of hospitalization *ρ* and the rate at which individuals are removed from the infectious population (*κ*), on the number of individuals in the infectious class *I*(*t*). The effect of increasing *ρ* is to reduce both the peak intensity and total infection burden by increasing the rate at which infectious individuals are removed from the infection pool. A lower value for the hospitalization saturation threshold *κ* will increase the speed of response of the system, and reduce the time required to contain the epidemic and lower its peak rate (skewness). In addition, in the combined intervention scenarios there was the greatest improvement in prevention due to the synergistic effect of the combination of interventions. This demonstrates that providing targeted control of clinical transition rates can significantly change how outbreaks develop, even when there are ongoing means of continuing transmission.

### 10.5 Sensitivity results: parameter ranking and intervention interpretation (PRCC/Sobol)

At last, we can evaluate how many input parameters really affect outputs of interest with some degree of uncertainty. This will be done using both the PRCC (Partial Rank Correlation Coefficient) method for monotonic (or near monotonic) relationships between inputs and their corresponding outputs and the Sobol index based variance-based methods when input-output relationships are highly nonlinear and/or have multiple interactions [23,24,25,26,27,28,29,30]. The sampling from a parameter’s admissible range is accomplished using Latin Hypercube Sampling. This provides an efficient stratification of the population; as a result, this method works well in moderate-to-high dimensions [30]. For every sampled value of parameters, an output vector which is a consistent measure of the response of the model, will be created. For example, with the baseline using the *R*_0_, *I*_max_, *t*_max_, *D*(*T*) & attack rates proxies, we generate PRCC (Partial Rank Correlation Coefficients) Heatmaps and Sobol Bar Plots [27,28,29,30]. The interpretation is explicitly related to a governmental agency (as opposed to a private-sector agency): specific factors associated with transmission limitation or routing into isolation at the cost of diverting from standard care are identified as parameters when they have an effect on the variability in severe outcomes, but when separated from spillover effects of highly variable *R*_0_ values have little effect on *D*(*T*). By answering the question “What are the specific mechanisms that remain important when there is uncertainty,” this study addresses an aspect of resilience (the Ulam-Hyers stability) through an evaluation of the different pathways of uncertainty to outcomes.

**Figure 7.**
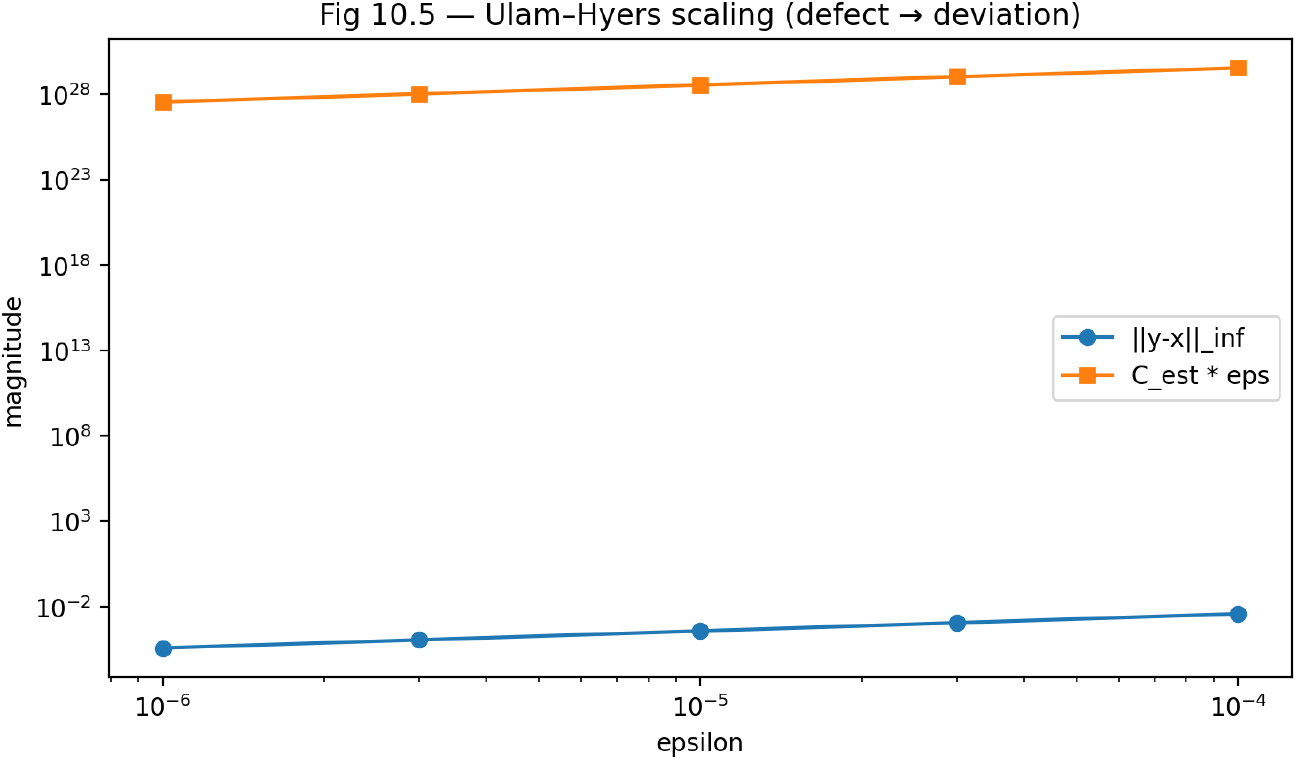
Comparison of observed deviation ∥*y* − *x*∥ _∞_ and theoretically calculated bound estimate *C*_est_ *ε* Testing of Ulam-Hyers Robustness through controlled defect injection has shown that Ulam-Hyers provides an upperbound on the accuracy of a solution, but does not guarantee that the error between the two solutions will remain constant. The steady state increase in size of both models has shown, by empirical methods of determining deviation from the original state, that the solutions to the measurements of the effect of the inject defect have a proportional response. Thus, since the numerical deviation from either of the model curves is always relatively lower than the theoretical prediction, we conclude that the fractional-order SEIHRD system is robust to perturbations (both in terms of operational performance) which also supports the validity of the model in relation to sensitivity analyses in that variability in parameters will not be hidden due to numerical instability.

## 11 Discussion

Human-to-human amplification and exogenous spillover forcing (recurrent seeding into exposed individuals) have been separated in this paper, helping to define their respective roles and how they influence the basic reproduction number, *R*_0_, which remains as a meaningful measurement of invasion capacity but changes based on context. The baseline *R*_0_ of an autonomous infection (*η*(*t*) = 0) is used to develop a new threshold for measuring invasion capacity; specifically that *R*_0_ < 1, or small infected initial perturbations will not be able to maintain their growth; and *R*_0_ > 1 indicates local instability and may allow outbreak amplification. A “disease-free” state does not exist when *η*(*t*) ≠ 0. The correct interpretation of the system forced with the presence of disease is that the same within-population transmission mechanism (captured by *R*_0_) remains in effect and is now sustained by continuous external introduction of disease into the population. Conclusions should be drawn looking at the relative difference between scenarios (different forcing intensities, different seasonality patterns of forcing, and/or any impulsive bursts of forcing), rather than through statements of absolute disease eradication. This view is consistent with the existing threshold logic regarding how the next generation will be constructed as well as the difference between the endogenous (self-generated) transmission of information and the exogenous (outside of one’s control) driving forces behind it. [18-19-26] Because of this second component, the fractional memory regime creates a second axis influencing policy interpretations. There are two components to the Caputo dynamic. First, *α* is a fractional exponent that creates a new scale for measuring time, and second, changing *α* results in changes in how previous states influence the evolution of current states through the memory kernel. As a consequence, the same set of nominal parameters can give rise to substantively different peak timing, peak widths, and tail persistence for *α* even for the same instantaneous vector fields. Such an observation has important implications for the face validity of claims exposed to decision-making scenarios: an intervention is deemed effective at a certain memory level (e.g., peak suppression) but can indeed appear as a peak delay or redistribution of the burden when examined from another memory perspective. The “scientific consensus” about this phenomenon would be that it works as a feature in function of the given memory regime. The fractional epidemic modeling studies have repeatedly stressed that memory can significantly affect the dynamics of an outbreak in a manner that cannot be accounted for by their integer-order counterparts. Therefore, *α*-robust statements are not just nice-to-have, but a necessity, [14,15,16]. In this context, Ulam-Hyers’ notion of stability adds a level of robustness that connects analysis and computation. The Ulam-Hyers’ method replaces the vague argument of “the simulation looks right” with a precise argument: “Given that the approximate trajectory has uniformly small defect in the equation being simulated, the approximate trajectory is uniformly close to the exact solution.” This is particularly important in fractional systems, as long memory can allow small defects to build up and result in global drifts. The generalized extension of the U-H theorem also supports nonlinear defect classes, as a more realistic modelling process involves the accumulation of sources of uncertainty, such as noise, discretization, and perturbations of parameters. From an operational point of view, U-H bounds provide a justification for defect/residual monitoring as a credibility check, not only computing a trajectory, but also quantifying whether or not this trajectory is guaranteed to be close, within certain constants, to a solution that is consistent with the model [4,10,31]. An alternative to U-H robustness can be obtained via a sensitivity analysis, where a different estimate is provided, namely what are the inputs that have the greatest influence on the results in terms of decision-relevant outputs over reasonable parameter ranges? Local derivative-based indices can also provide useful information if the model has an analytically tractable expression of *R*_0_, since results can then be obtained immediately in the baseline case. Global methods, which include structured sampling of admissible ranges (Latin Hyper-Cube sampling), PRCC in monotone regimes, as well as variance-based Sobol indices in interaction-dominated regimes, are used to determine which of the following factors dominates the burden of severe outcomes: hospitalization load or cumulative deaths) are primarily driven by community transmission, care-setting amplification, routing/isolation levers, or spillover intensity. The payoff is that it results in a disciplined prioritization logic in which parameters that are of greatest importance for severe outcome under uncertainty can be considered for intervention, and parameters that affect outcome of invasion but are not important for severe outcome under persistent spillover should be approached cautiously as a mechanistic rather than an intervention parameter, [27,30]. There are a number of limitations that specify the range of inference. For example, the data streams from the Nipah infections can have sparsity and heterogeneity. This leads to a weak identifiability of the models, and different parameters can be considered as a good fit over a small time window. Second, the deterministic nature of the mean-field approach eliminates the diversity of contacts and environments, which could be a source of bias for outcome mapping if superspreading events are important. Third, spillover is necessarily given a stylized form as a forcing term; although this is appropriate in a scenario-based approach, the real ecological drivers of spillover can be complex and time-varying, possibly requiring covariates or stochastic structure to be represented appropriately. While these limitations do not affect the mechanistic separation of endogenous thresholding from exogenous seeding, they caution against the interpretation of the results, which are to be understood as conditional upon the observation operator, admissible forcing family, and mixing assumptions, [23,31,32].

## 12 Conclusion

This study developed and analyzed a Caputo fractional order SEIHRD model for the dynamics of Nipah with the consideration of the effects of spillovers as an explicit exogenous input to the model for the infected category. We have established the basic qualitative properties of a solution within a biologically meaningful invariant region, which are necessary for epidemiological reliability: the solution is positive, bounded, and depends continuously on initial conditions. The well-posedness was then established through a fixed point formulation based on the Volterra-type integral representation that is inherent to Caputo dynamics, and a compactness-based alternative when global contraction is not available. In relation to the problem of threshold dynamics, we have refined the natural definition of the disease-free equilibrium and the reproduction number for the autonomous underlying model with zero spillover, with the non-zero forcing being handled via scenario comparison instead of results for eradication. Besides traditional stability analysis, Ulam-Hyers stability, and Generalized Ulam-Hyers stability, we added an explicit robustness layer, where uniformly small defects, such as from discretization, inconsistent measurement, or mild model variation, correspond to uniformly controlled deviation in the trajectories, where the control values can be interpreted in terms of memory order, time horizon, and Lipschitz amplification. The sensitivity and uncertainty quantification aspects complemented this notion of robustness by indicating which inputs are most significant for certain aspects of the decision over reasonable parameter ranges. Finally, we presented a predictor-corrector Adams-type scheme tailored for the Caputo equation along with some practical verification and reproducibility features (refinement tests, defect monitoring, positivity/invariant set verification, and machine-readable configuration) so that the conclusions drawn can be audited rather than assumed. Possible extensions of this work may be the development of variable order memory models that can accommodate changing patterns of behavior and clinical data, as well as stochastic/covariate driven spillover forcing, contact structure models beyond the use of mean field mixing, and the development of Bayesian methods for calibration/data assimilation to propagate uncertainty through observations to the ranges of interest.

## Future Work

Several natural extensions of the present study are worth pursuing. From a numerical analysis point of view, the construction and analysis of fractional differential shift integrators for Caputo-type initial value problems is an interesting area, especially with regard to accuracy, stability, and computational complexity. This type of integrator can offer a flexible framework for long-memory systems, further improving the reliability of large-scale epidemic simulations [33, 35]. On the modeling side, extensions of the current framework towards classical and fractional SIR-type structures, including more elaborate numerical experimentation, can offer the possibility of directly comparing deterministic and memory-driven epidemic dynamics. In particular, fractional order solver usage in concert with transparent simulation pipelines has been identified as a potential improvement for model interpretability and reproducibility in infectious diseases [34]. Other than the epidemiological applications, the underlying fractional shift operator has more general applicability in information processing and security-oriented dynamics. Recent advances in Caputo-based fractional encryption mechanisms imply that similar operator theory-based ideas may be utilized for the development of robust dynamical systems with sensitivity[33,35]. Finally, the integration of fractional order models with early warning signs based on data-driven indicators, such as EWMA-based surveillance indices using wastewater or population data, is an interesting area of interdisciplinary research. The integration may potentially close the gap between modeling and real-time public health monitoring, hence providing the way forward towards predictive and adaptive epidemic response strategies [36].

## Data Availability

All data produced in the present study are contained within the manuscript.

